# Skeletal effects of sleeve gastrectomy, by sex and menopausal status and in comparison to Roux-en-Y gastric bypass surgery

**DOI:** 10.1101/2024.06.25.24309368

**Authors:** Karin C Wu, Po-Hung Wu, Galateia Kazakia, Sheena Patel, Dennis M Black, Thomas F Lang, Tiffany Y Kim, Nicole J King, Thomas J. Hoffman, Hanling Chang, Gaia Linfield, Sarah Palilla, Stanley J Rogers, Jonathan T Carter, Andrew M Posselt, Anne L Schafer

## Abstract

**Context:** Roux-en-Y gastric bypass (RYGB) has deleterious effects on bone mass, microarchitecture, and strength. Data are lacking on the skeletal effects of sleeve gastrectomy (SG), now the most commonly performed bariatric surgical procedure.

**Objective:** We examined changes in bone turnover, areal and volumetric bone mineral density (aBMD, vBMD), and appendicular bone microarchitecture and estimated strength after SG. We compared the results to those previously reported after RYGB, hypothesizing lesser effects after SG than RYGB.

**Design, Setting, Participants:** Prospective observational cohort study of 54 adults with obesity undergoing SG at an academic center.

**Main Outcome Measure(s):** Skeletal characterization with biochemical markers of bone turnover, dual-energy X-ray absorptiometry (DXA), quantitative computed tomography (QCT), and high-resolution peripheral QCT (HR-pQCT) was performed preoperatively and 6- and 12-months postoperatively.

**Results:** Over 12 months, mean percentage weight loss was 28.8%. Bone turnover marker levels increased, and total hip aBMD decreased −8.0% (95% CI −9.1%, −6.7%, p<0.01). Spinal aBMD and vBMD declines were larger in postmenopausal women than men. Tibial and radial trabecular and cortical microstructure worsened, as did tibial estimated strength, particularly in postmenopausal women. When compared to data from a RYGB cohort with identical design and measurements, some SG biochemical, vBMD, and radial microstructural parameters were smaller, while other changes were not.

**Conclusions:** Bone mass, microstructure, and strength decrease after SG. Some skeletal parameters change less after SG than after RYGB, while for others, we find no evidence for smaller effects after SG. Postmenopausal women may be at highest risk of skeletal consequences after SG.

## Introduction

One in 11 US adults (9.2%), and one in nine US women (11.2%), are living with severe or Class 3 obesity (body mass index [BMI] ≥ 40 kg/m^2^) (1). Obesity is linked to multiple comorbid conditions and is associated with increased mortality (2, 3). Metabolic and bariatric surgery (MBS) is a highly effective intervention for severe obesity, leading to marked and durable weight loss, improvement in obesity-associated diseases, and decreased mortality (4, 5). However, Roux-en-Y gastric bypass (RYGB), previously the most popular procedure and often considered the gold-standard for metabolic benefit, has been shown to induce detrimental effects on calcium homeostasis and skeletal health, with increases in bone turnover, decreases in bone mineral density (BMD), weakened bone microarchitecture, and increased fracture incidence (6–15).

Sleeve gastrectomy (SG) has emerged over the last decade and has overtaken RYGB as the most commonly performed weight loss operation in the US and around the world (16, 17). SG involves the removal of 60-80% of the stomach mass along the greater curvature, without the alteration of the intestinal pathway that characterizes RYGB. Its rise in patient and surgeon preference is largely due to the simpler surgical approach, lower peri- and postoperative complication and mortality rates, and roughly clinically comparable long-term weight loss and metabolic benefits (5, 18, 19). However, the impact of SG on skeletal health is still not well characterized.

Published studies of skeletal effects of SG have generally examined areal BMD (aBMD) by dual-energy X-ray absorptiometry (DXA), but DXA may be biased in the setting of marked body composition changes and degenerative changes in the spine. Quantitative computed tomography (QCT) and high-resolution peripheral QCT (HR-pQCT) are three-dimensional, non-invasive methods for assessing volumetric BMD (vBMD) at the axial and appendicular skeleton and for examining cortical and trabecular bone microarchitecture and estimated bone strength (20, 21). Further, studies have inconsistently addressed the vitamin D deficiency that is so common in severe obesity and with dietary restriction. Moreover, the relative skeletal effects of SG by sex and menopausal status are uncertain. Because the majority of individuals who undergo bariatric surgery are premenopausal women, studies of postoperative skeletal changes after SG have largely focused on that group (22–25). However, postmenopausal women’s skeletons, already vulnerable due to age and sex steroid deficiency, may be particularly affected, as we have shown after RYGB (13). If this is the case after SG, there may be clinical implications for targeted screening and postoperative management.

The aim of this study was to examine the effects of SG on aBMD, vBMD, and appendicular microarchitecture and estimated strength in a cohort of pre- and postmenopausal women and men with severe obesity, in the setting of optimized 25-hydroxyvitamin D (25OHD) status and recommended calcium intake. We also aimed to compare observed skeletal changes after SG with changes reported in our previous cohort study of RYGB (13), which employed an identical protocol and measurements. We hypothesized that bone mass, microstructure, and estimated strength would decline, but to a lesser extent than RYGB, after SG.

## Materials and Methods

### Study design & population

We recruited women and men with severe obesity, aged 25 to 70 years, from an academic bariatric surgery center (the University of California, San Francisco [UCSF]), as previously described (26). Participants were eligible if they were scheduled for an upcoming SG procedure, which was done laparoscopically and sometimes with robotic assist. Eligibility for bariatric surgery at our institution was in accordance with the 1991 National Institutes of Health consensus conference (BMI ≥ 40 kg/m^2^ or ≥ 35 kg/m^2^ with obesity-related comorbid conditions) (27) and failure to lose weight with medical management. For this cohort study, we excluded perimenopausal women (last menses > 3 months but < 4 years ago), and additional exclusion criteria included a history of intestinal malabsorption or prior bariatric surgery, use of medications known to impact bone and mineral metabolism (*e.g.* osteoporosis pharmacotherapy, glucocorticoids, thiazolidinediones, aromatase inhibitors, androgen deprivation therapy), diseases known to affect bone (*e.g.* primary hyperparathyroidism, Paget’s disease, hyperthyroidism defined by TSH < 0.1 mIU/L, clinically significant liver disease), illicit drug use or alcohol use > 3 drinks/day, hypercalcemia (serum calcium > 10.2 mg/dL), chronic kidney disease stage 4 or lower (eGFR < 30 mL/min/1.73m^2^), and weight > 200 kg (weight limit of the DXA scanner).

Participants underwent study measurements preoperatively (no more than 3 months before the SG) and at 6 months and 12 months postoperatively. However, the COVID19 pandemic impeded our ability to adhere to the predefined timeline, particularly for the 12-month postoperative visit. The mean ± standard deviation (SD) for the 6-month postoperative timepoint was 7.5 ± 1.6 months and for the 12-month postoperative timepoint was 17.6 ± 9.7 months. After examining the distribution, we excluded data collected after 20 months postoperatively.

Protocol approval was obtained from the Institutional Review Board at the University of California, San Francisco. Written informed consent was obtained from all participants. The study was registered at the US National Institutes of Health (ClinicalTrials.gov, NCT02778490).

For comparison between SG and RYGB procedures, we used a previously completed (5 years prior) pre-post observational cohort study of 48 women and men ages 25 to 70 years with severe obesity undergoing RYGB at UCSF and the San Francisco Veterans Affairs Health Care System (13, 28). The two cohort studies employed the identical approach/protocol, and measurements and were made by the same research team, sequentially. The SG study was designed to accommodate careful comparisons between these two cohorts.

### Calcium and vitamin D supplementation

The research protocol included standardization of calcium intake and attention to vitamin D status, following emerging clinical practice guidelines (29, 30). Vitamin D and chewable calcium citrate supplements were provided upon enrollment (at least 2 weeks prior to the preoperative measurements) and supplied throughout the study period. At enrollment, low 25OHD levels were repleted to a target level ≥ 30 ng/mL, and vitamin D was supplemented with at least 3000 IU/day. Each participant’s total daily calcium intake was brought to 1200 mg through individualized calcium citrate dosing, based on estimation of intake from a validated screener (31). Postoperatively, 25OHD levels and estimated dietary calcium intake were monitored, and each participant’s supplement doses were adjusted to maintain the vitamin D and calcium intake goals.

### Study measures

#### DXA bone density and body composition

Areal BMD (aBMD, g/cm^2^) at the lumbar spine (L1 to L4), proximal femur, and distal radius was measured using DXA (Horizon A, Hologic) at each study time point. Whole-body DXA were also performed for assessment of estimated total and regional body composition, including percentage body fat (32). If the body dimensions exceeded the width of the scanning area, the DXA manufacturer’s reflection technique was utilized.

#### QCT for spine bone density

Spinal volumetric BMD (vBMD, g/cm^3^) was assessed by QCT at the L3 and L4 vertebrae at 120 kVp, 200 mAs (General Electric’s VCT64 scanner; General Electric, Milwaukee, WI, USA) at each study time point and was analyzed according to methods previously described (Mindways Software, Austin, TX, USA) (33, 34).

#### HRpQCT for appendicular bone density, microarchitecture, and estimated strength

Participants were imaged with a HRpQCT system (XtemeCT1, Scanco Medical, Brüttisellen, Switzerland) at each study time point, using the manufacturer’s standard in vivo protocol (source potential 60 kVp, tube current 900 μA, isotropic 82 μm nominal resolution) (35–37). The nondominant forearm and ankle were scanned. Fixed scan regions started at 9.5 mm proximal to the mid-joint line for the ultradistal radius and 22.5 mm for the ultradistal tibia and extended proximally for 9.02 mm (110 slices). Images were individually examined and rated for artifacts as a result of soft tissue extension outside the field of view.

HR-pQCT images were analyzed using the manufacturer’s standard clinical evaluation protocol in Image Processing Language (IPL v5.08b, Scanco Medical) (38–40). Contours identifying the periosteal perimeter of the bone were drawn semi-automatically using an edge-finding algorithm (40) and manually examined and modified as necessary. A threshold-based process was used to segment cortical and trabecular regions (40) and was manually checked for accurate segmentation. Trabecular structure and cortical parameters were assessed using methods previously described (41). Linear elastic micro-finite element analysis (μFEA, Scanco FE Software, Scanco Medical) was used to calculate biochemical properties (41–43).

#### Other measures

Weight and height were measured at each study time point, and BMI (kg/m^2^) was calculated. Waist circumference was measured at the level directly below the lowest rib, and hip circumference at the maximum extension of the buttocks, viewed from the side.

Participants collected 24-hour urine specimens at each time point for measurement of urinary calcium and creatinine. Serum samples were collected after an overnight fast at each study time point. Basic chemistries including calcium, albumin, and phosphate, 25OHD, and parathyroid hormone (PTH) were measured at a commercial laboratory (Quest Diagnostics, Secaucus, NJ, USA). Additional serum samples were stored at −80°C until batch analyzed in a central laboratory (MaineHealth Institute for Research, Scarborough, ME, USA). Serum C-terminal cross-linked telopeptide (CTx; a marker of bone resorption), procollagen type 1 N-terminal propeptide (P1NP; a marker of bone formation), and 1,25-dihydroxyvitamin D (1,25[OH]_2_D) were measured by chemiluminescence on an auto-analyzer (iSYS, Immunodiagnostic Systems, Scottsdale, AZ). The inter- and intra-assay coefficients of variation (CV) were 6.0% and 3.2% for CTx, 5.0% and 2.9% for P1NP, and 11.1% and 6.4% for 1,25(OH)_2_D. Intestinal fractional calcium absorption (FCA) was measured preoperatively and at 6 months postoperatively by a gold-standard dual stable calcium isotope tracer method, as previously reported (26).

### Statistical methods

Sample size calculations for this study were based on the mean changes, SD, and covariance parameters identified in our previous pre-post RYGB cohort study (13), which had comparable study population, study protocol, and measurements. We calculated that a sample of 50 participants would provide 80% power in 2-sided tests with α = 0.05 to detect a percentage reduction in femoral neck, total hip, and lumbar spine aBMD of 2.0%, 2.1%, and 1.8%, respectively. These approximate the least significant change for DXA at our institution and the change commonly considered clinically significant.

Baseline descriptive data were reported as means ± SD, median (interquartile range [IQR]), or proportions (N [%]). One-way ANOVA or Kruskal-Wallis were used as appropriate to determine differences in baseline characteristics between sex and menopausal status (premenopausal women, postmenopausal women, and men). Changes in weight and bone parameters over time after SG were analyzed using mixed effect models with repeated measures with random subject intercept and slope and fixed effect of time, with and without the interactions of sex and menopausal status. Models were fitted with maximum likelihood estimation and were tested for non-linear relationships with a quadratic time term. The percentage change in bone parameters over 6 and 12 months and the confidence intervals for the predicted percentage changes were estimated with model-based parametric bootstrap with 1000 resamples. Participants were excluded from analysis if the 6-month postoperative percentage change for that bone parameter was > 3SD above or below the mean, suggesting a problem with the measurement of that parameter.

Comparisons of the changes in bone imaging parameters, body composition, and laboratory parameters between RYGB and SG were analyzed similarly using mixed effect models with repeated measures with random subject intercept and slope and fixed effect of time*procedure. These models were adjusted for age, sex, menopausal status, race, baseline weight, and diabetes status, which were determined a priori in light of the relevant biology. Paired t-test or Wilcoxon signed-rank tests were performed as appropriate to determine whether the changes in bone turnover markers and FCA between preoperative and 6-month postoperative timepoints differed between RYGB and SG cohorts. Linear models were then used to estimate the adjusted associations. The 6-month postoperative data was used for comparison because it is the study visit that is more closely matched in time since surgery between the two cohorts, given the impact of the COVID19 pandemic on follow-up after SG. Further, the use of one postoperative timepoint minimize the batch effect of serum analyses, and FCA was measured only at the 6-month postoperative timepoint.

Statistical significance level was set at a two-sided p-value < 0.05, with corresponding 95% confidence intervals. All data sets were managed using SAS software (version 9.4, SAS Institute Inc., Cary, North Carolina, USA) and all statistical analyses were done using R (version 4.0.3) (44).

## Results

### Baseline SG participant characteristics

Fifty-four participants completed both pre- and postoperative skeletal imaging assessments. Preoperatively, mean age ± SD was 46.2 ± 11.1 years, weight was 126.0 ± 23.4 kg, and BMI was 45.2 ± 7.5 kg/m^2^ (Table 1). Of the 54 participants, 41 (76%) were women, with 15 (37%) of those postmenopausal and 26 (63%) premenopausal. Mean 25OHD level at the preoperative skeletal imaging assessments was 40.6 ± 13.7 ng/mL, which reflected preoperative repletion of low levels that had been performed in accordance with clinical practice guidelines (29, 30). Mean serum calcium, PTH, 1,25(OH)_2_D, serum creatinine, and serum phosphorate were within their reference ranges. Median 24-hour urinary calcium level was 172 mg (IQR 80 to 270 mg).

**Table 1.**
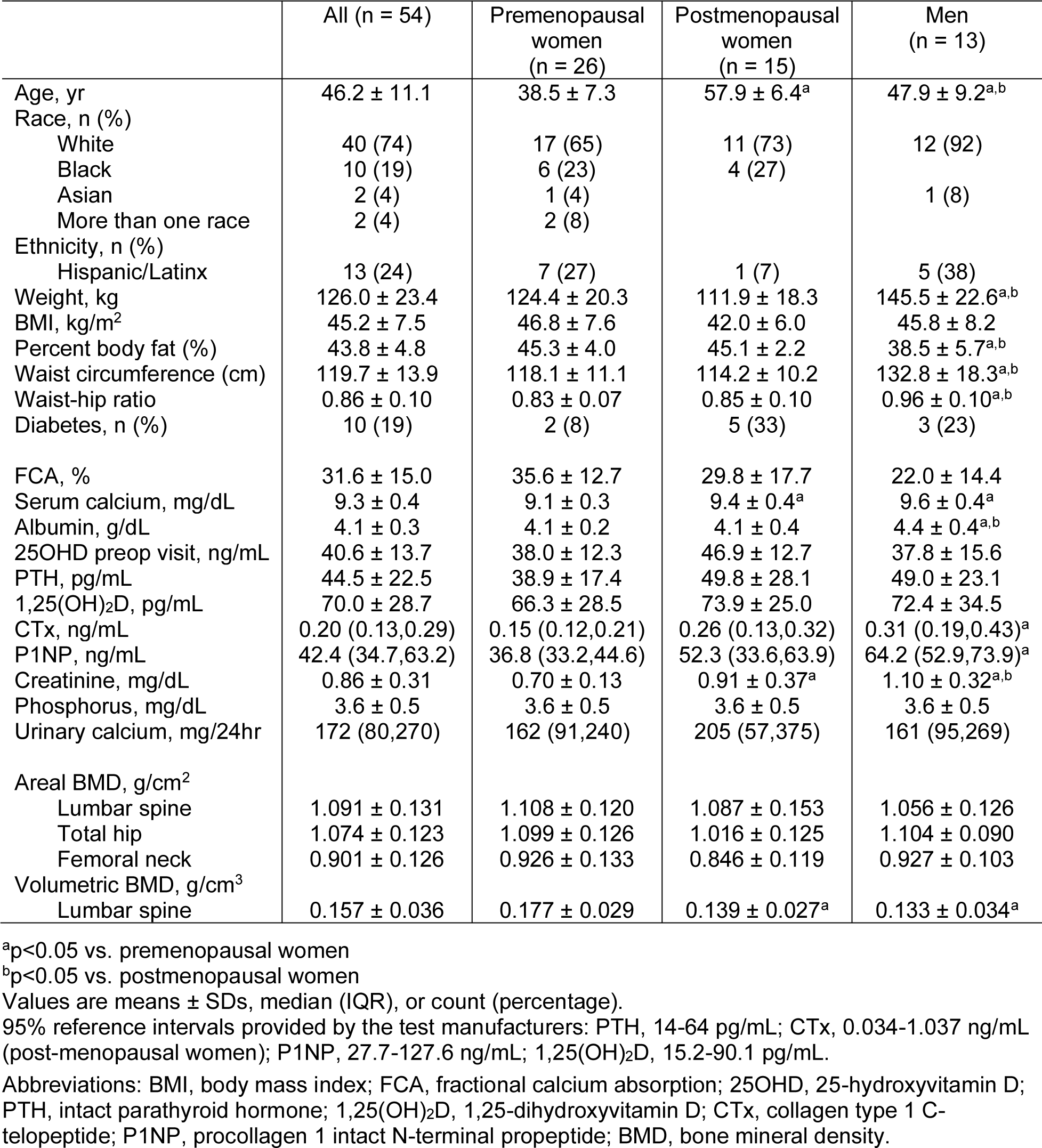
Baseline characteristics of SG participants.

|  | All (n = 54) | Premenopausal women (n = 26) | Postmenopausal women (n = 15) | Men (n = 13) |
| --- | --- | --- | --- | --- |
| Age, yr | 46.2 ± 11.1 | 38.5 ± 7.3 | 57.9 ± 6.4<sup>a</sup> | 47.9 ± 9.2<sup>a,b</sup> |
| Race, n (%) |  |  |  |  |
| White | 40 (74) | 17 (65) | 11 (73) | 12 (92) |
| Black | 10 (19) | 6 (23) | 4 (27) |  |
| Asian | 2 (4) | 1 (4) |  | 1 (8) |
| More than one race | 2 (4) | 2 (8) |  |  |
| Ethnicity, n (%) |  |  |  |  |
| Hispanic/Latinx | 13 (24) | 7 (27) | 1 (7) | 5 (38) |
| Weight, kg | 126.0 ± 23.4 | 124.4 ± 20.3 | 111.9 ± 18.3 | 145.5 ± 22.6<sup>a,b</sup> |
| BMI, kg/m<sup>2</sup> | 45.2 ± 7.5 | 46.8 ± 7.6 | 42.0 ± 6.0 | 45.8 ± 8.2 |
| Percent body fat (%) | 43.8 ± 4.8 | 45.3 ± 4.0 | 45.1 ± 2.2 | 38.5 ± 5.7<sup>a,b</sup> |
| Waist circumference (cm) | 119.7 ± 13.9 | 118.1 ± 11.1 | 114.2 ± 10.2 | 132.8 ± 18.3<sup>a,b</sup> |
| Waist-hip ratio | 0.86 ± 0.10 | 0.83 ± 0.07 | 0.85 ± 0.10 | 0.96 ± 0.10<sup>a,b</sup> |
| Diabetes, n (%) | 10 (19) | 2 (8) | 5 (33) | 3 (23) |
| FCA, % | 31.6 ± 15.0 | 35.6 ± 12.7 | 29.8 ± 17.7 | 22.0 ± 14.4 |
| Serum calcium, mg/dL | 9.3 ± 0.4 | 9.1 ± 0.3 | 9.4 ± 0.4<sup>a</sup> | 9.6 ± 0.4<sup>a</sup> |
| Albumin, g/dL | 4.1 ± 0.3 | 4.1 ± 0.2 | 4.1 ± 0.4 | 4.4 ± 0.4<sup>a,b</sup> |
| 25OHD preop visit, ng/mL | 40.6 ± 13.7 | 38.0 ± 12.3 | 46.9 ± 12.7 | 37.8 ± 15.6 |
| PTH, pg/mL | 44.5 ± 22.5 | 38.9 ± 17.4 | 49.8 ± 28.1 | 49.0 ± 23.1 |
| 1,25(OH)<sub>2</sub>D, pg/mL | 70.0 ± 28.7 | 66.3 ± 28.5 | 73.9 ± 25.0 | 72.4 ± 34.5 |
| CTx, ng/mL | 0.20 (0.13,0.29) | 0.15 (0.12,0.21) | 0.26 (0.13,0.32) | 0.31 (0.19,0.43)<sup>a</sup> |
| P1NP, ng/mL | 42.4 (34.7,63.2) | 36.8 (33.2,44.6) | 52.3 (33.6,63.9) | 64.2 (52.9,73.9)<sup>a</sup> |
| Creatinine, mg/dL | 0.86 ± 0.31 | 0.70 ± 0.13 | 0.91 ± 0.37<sup>a</sup> | 1.10 ± 0.32<sup>a,b</sup> |
| Phosphorus, mg/dL | 3.6 ± 0.5 | 3.6 ± 0.5 | 3.6 ± 0.5 | 3.6 ± 0.5 |
| Urinary calcium, mg/24hr | 172 (80,270) | 162 (91,240) | 205 (57,375) | 161 (95,269) |
| Areal BMD, g/cm<sup>2</sup> |  |  |  |  |
| Lumbar spine | 1.091 ± 0.131 | 1.108 ± 0.120 | 1.087 ± 0.153 | 1.056 ± 0.126 |
| Total hip | 1.074 ± 0.123 | 1.099 ± 0.126 | 1.016 ± 0.125 | 1.104 ± 0.090 |
| Femoral neck | 0.901 ± 0.126 | 0.926 ± 0.133 | 0.846 ± 0.119 | 0.927 ± 0.103 |
| Volumetric BMD, g/cm<sup>3</sup> |  |  |  |  |
| Lumbar spine | 0.157 ± 0.036 | 0.177 ± 0.029 | 0.139 ± 0.027<sup>a</sup> | 0.133 ± 0.034<sup>a</sup> |

<sup>a</sup>p<0.05 vs. premenopausal women<sup>b</sup>p<0.05 vs. postmenopausal women
Values are means ± SDs, median (IQR), or count (percentage).
95% reference intervals provided by the test manufacturers: PTH, 14-64 pg/mL; CTx, 0.034-1.037 ng/mL (post-menopausal women); P1NP, 27.7-127.6 ng/mL; 1,25(OH)<sub>2</sub>D, 15.2-90.1 pg/mL.Abbreviations: BMI, body mass index; FCA, fractional calcium absorption; 25OHD, 25-hydroxyvitamin D; PTH, intact parathyroid hormone; 1,25(OH)<sub>2</sub>D, 1,25-dihydroxyvitamin D; CTx, collagen type 1 C-telopeptide; P1NP, procollagen 1 intact N-terminal propeptide; BMD, bone mineral density.

Mean age for men was higher than for premenopausal women, but lower than for postmenopausal women. Preoperative BMI was similar across sex and menopause groups. However, men had higher preoperative weight, lower percentage body fat, higher waist circumference, and higher waist-to-hip ratio. Vitamin D status, PTH, and other measures of calcium homeostasis (FCA and 24-hour urinary calcium) did not differ by sex and menopausal status. Men had higher bone turnover marker levels compared to premenopausal women. Baseline aBMD by DXA at the spine and proximal femur did not differ by sex and menopause status. Postmenopausal women and men had lower preoperative lumbar spine vBMD than premenopausal women.

### Weight loss after SG

Weight loss was dramatic after SG, with predicted 6-month percentage weight loss −21.8% (95% CI −23.8%, −19.9%) of preoperative weight, and 12-month percentage weight loss −28.8% (−31.3%, −26.0%) (p < 0.01 for both). Percentage weight loss did not differ by sex or menopausal status.

### Changes in biochemical markers of bone turnover after SG

Bone turnover markers increased markedly after SG. Serum CTX, a marker of bone resorption, increased by a median +156% (IQR +89% to +279%) over 6 months, then at 12 months it was still +87% above baseline (IQR +38% to +175%) (p < 0.01 for both). Serum P1NP, a marker of bone formation, increased by a median +58% (IQR +33% to +100%) over 6 months, then at 12 months it was still +47% above baseline (IQR +11% to +82%) (p < 0.01 for both).

### Changes in BMD after SG

Areal BMD at the proximal femur (DXA) decreased progressively after SG (Table 2), with predicted 6- and 12-month declines at the femoral neck of −3.4% and −6.7%, respectively, and at the total hip of −5.4% and −8.0%, respectively (p<0.01 for both). There was a decline in lumbar spine aBMD (p = 0.03) and a trend for decline in lumbar spine vBMD (p = 0.07). The decline in lumbar spine aBMD by DXA was larger for postmenopausal women than for premenopausal women or men (Figure 1a). The decline in lumbar spine vBMD by QCT was larger for women than for men, with a decline of −5.8% in premenopausal women and −3.6% in postmenopausal women vs. an increase of +8.2% in men over 12 months (Figure 1b).

**Figure 1:**
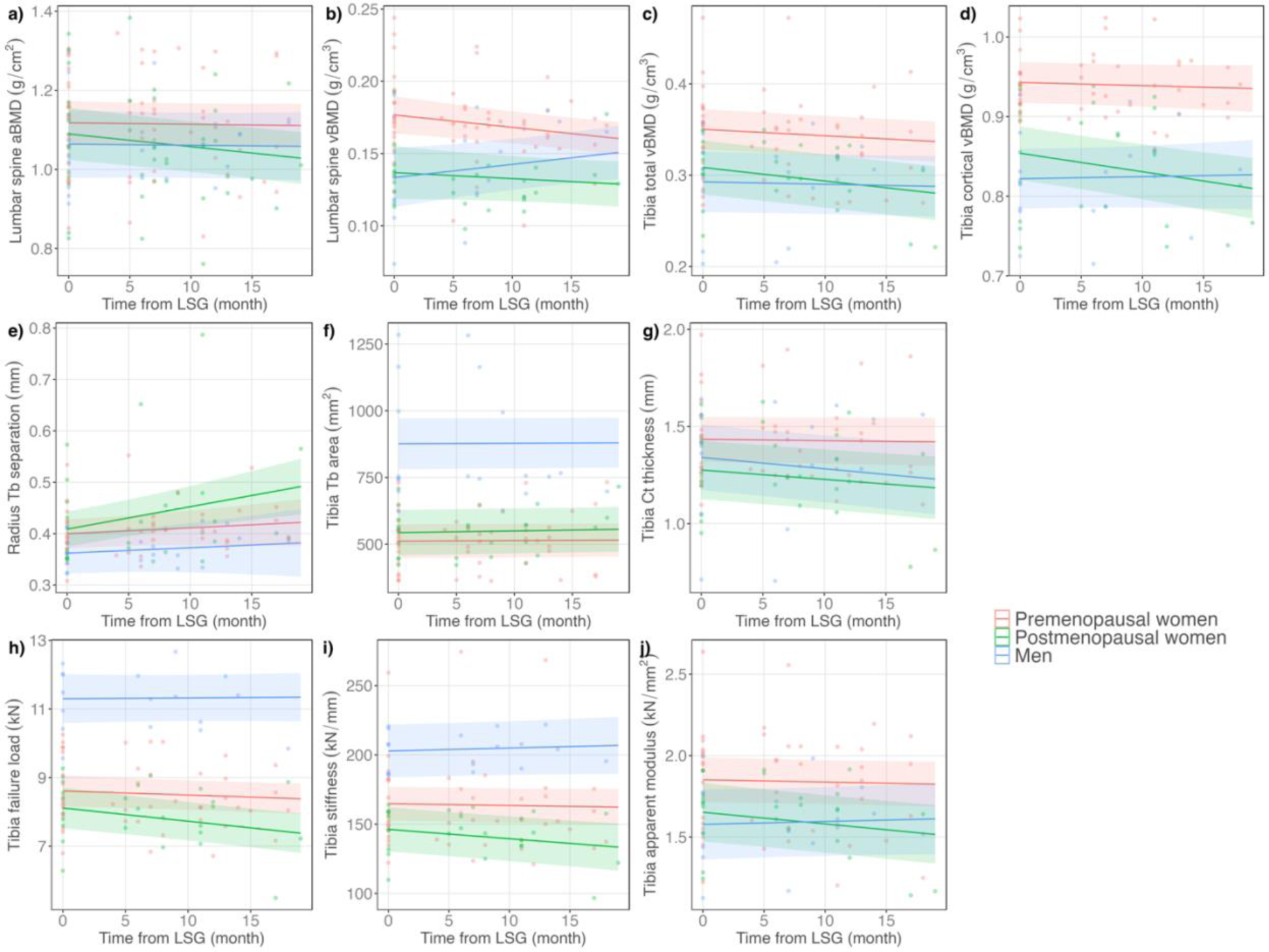
Changes in BMD, bone microarchitecture, and estimated strength that were significantly different by sex and menopausal status. Shaded areas represent the 95% confidence intervals.

**Table 2.** Predicted percentage changes in BMD after SG.

|  | 6-month % change | 12-month % change | *p* |
| --- | --- | --- | --- |
| *DXA* (n = 49) |  |  |  |
| Total hip aBMD (g/cm<sup>2</sup>) | -5.4 [-6.4,-4.3] | -8.0 [-9.1,-6.7] | <0.01 |
| Femoral neck aBMD (g/cm<sup>2</sup>) | -3.4 [-4.2,-2.5] | -6.7 [-8.3,-5.2] | <0.01 |
| Lumbar spine aBMD (g/cm<sup>2</sup>) | -0.7 [-1.4,-0.1] | -1.4 [-2.6,-0.3] | 0.03 |
| Distal 1/3 radius aBMD (g/cm<sup>2</sup>) | +0.3 [-0.1,0.7] | +0.7 [-0.1,1.6] | 0.11 |
| *QCT* (n = 38) |  |  |  |
| Lumbar spine vBMD (g/cm<sup>3</sup>) | -1.6 [-2.9,-0.1] | -3.1 [-6.1,0.1] | 0.07 |
| *HRpQCT* (n = 37) |  |  |  |
| Radius total vBMD (g/cm<sup>3</sup>) | -0.4 [-1.1,0.2] | -0.8 [-2.2,0.4] | 0.22 |
| Radius trabecular vBMD (g/cm<sup>3</sup>) | -0.9 [-1.6,-0.2] | -1.9 [-3.3,-0.5] | 0.01 |
| Radius cortical vBMD (g/cm<sup>3</sup>) | +0.2 [-0.1,0.4] | 0.4 [-0.1,0.9] | 0.17 |
| Tibia total vBMD (g/cm<sup>3</sup>) | -1.6 [-2.2,-1.0] | -3.1 [-4.2,-1.9] | <0.01 |
| Tibia trabecular vBMD (g/cm<sup>3</sup>) | -1.1 [-1.7,-0.4] | -2.1 [-3.3,-0.8] | <0.01 |
| Tibia cortical vBMD (g/cm<sup>3</sup>) | +0.0 [-0.6,0.7] | -1.0 [-2.2,0.1] | 0.20 |

Values are model predictions ± 95%CI
P-values, which compare to no change, were calculated with mixed effect models with repeated measures with random participant intercept and slope and fixed effect of time.
Abbreviations: DXA, dual-energy X-ray absorptiometry; aBMD, areal bone mineral density; QCT, quantitative computed tomography; vBMD, volumetric bone mineral density; HRpQCT, high-resolution peripheral quantitative computed tomography.

Declines in appendicular vBMD after SG, measured by HR-pQCT, were smaller in magnitude than declines in axial BMD. The decline in trabecular vBMD at the radius and tibia and total vBMD at the tibia were statistically significant. The declines in total vBMD and cortical vBMD at the tibia were largest in postmenopausal women (tibia total vBMD over 12 months: −5.8% in postmenopausal women vs. −2.4% in premenopausal women vs. −1.1% in men, Figure 1c; tibia cortical vBMD over 12 months: −3.3% in postmenopausal women vs. −0.5% in premenopausal women vs. +0.4% in men, Figure 1d).

### Changes in bone microarchitecture and estimated strength after SG

Within the trabecular compartment, changes in microarchitecture associated with diminished skeletal strength were observed at the tibia and the radius (Table 3). There were decreases in trabecular number and increases in trabecular separation and trabecular heterogeneity, as well as in trabecular thickness. Increases in trabecular separation at the radius and trabecular area at the tibia were larger for postmenopausal women (Figures 1e, 1f). Within the cortical compartment, cortical pore size increased at both the radius and tibia (Table 3). There was also a decrease in cortical thickness at the tibia, which was larger for postmenopausal women than premenopausal women (Figure 1g).

**Table 3.** Predicted percentage changes in bone microarchitecture and estimated strength after SG.

| HRpQCT (n = 37) | 6-month % change | 12-month % change | p |
| --- | --- | --- | --- |
| *Bone microarchitecture* |  |  |  |
| Radius |  |  |  |
| Trabecular area (mm<sup>2</sup>) | -0.1 [-0.3,0.2] | -0.1 [-0.6,0.3] | 0.64 |
| Trabecular number (per mm) | -2.8 [-4.0,-1.6] | -5.6 [-8.0,-3.3] | <0.01 |
| Trabecular separation (mm) | +3.3 [1.1,5.4] | +6.6 [2.6,11.0] | <0.01 |
| Trabecular thickness (mm) | +1.7 [0.1,3.4] | +3.3 [0.1,6.3] | 0.04 |
| Trabecular heterogeneity (per mm) | +7.6 [0.7,14.4] | +15.2 [0.5,29.4] | 0.05 |
| Cortical thickness (mm) | -0.1 [-0.7,0.6] | -0.2 [-1.5,1.1] | 0.77 |
| Cortical porosity (%) | -0.3 [-4.0,3.7] | -0.5 [-7.7,7.4] | 0.90 |
| Cortical pore size (mm) | +1.8 [0.7,2.9] | +3.5 [1.4,5.6] | <0.01 |
| Tibia |  |  |  |
| Trabecular area (mm<sup>2</sup>) | +0.3 [0.2,0.5] | +0.7 [0.4,1.0] | <0.01 |
| Trabecular number (per mm) | -4.2 [-5.3,-2.9] | -8.3 [-10.7,-5.9] | <0.01 |
| Trabecular separation (mm) | +5.0 [3.2,6.7] | +10.0 [6.2,13.6] | <0.01 |
| Trabecular thickness (mm) | +3.1 [1.4,5.0] | +6.3 [2.6,9.8] | <0.01 |
| Trabecular heterogeneity (per mm) | +6.6 [4.4,8.9] | +13.2 [8.4,17.3] | <0.01 |
| Cortical thickness (mm) | -1.3 [-2.1,-0.6] | -2.6 [-4.2,-1.1] | <0.01 |
| Cortical porosity (%) | +2.7 [-2.0,7.7] | +5.5 [-4.0,15.3] | 0.24 |
| Cortical pore size (mm) | +2.7 [1.7,3.7] | +5.4 [3.3,7.4] | <0.01 |
| *Estimated strength* |  |  |  |
| Radius |  |  |  |
| Failure load (kN) | -0.5 [-1.2,0.3] | -0.9 [-2.4,0.5] | 0.20 |
| Stiffness (kN/mm) | -0.1 [-1.1,0.8] | -0.3 [-2.2,1.6] | 0.77 |
| Apparent modulus (kN/mm<sup>2</sup>) | -0.3 [-1.2,0.5] | -0.7 [-2.2,1.1] | 0.44 |
| Tibia |  |  |  |
| Failure load (kN) | -1.1 [-1.9,-0.3] | -2.3 [-3.9,-0.7] | <0.01 |
| Stiffness (kN/mm) | -0.8 [-1.7,0.1] | -1.7 [-3.4,-0.1] | 0.06 |
| Apparent modulus (kN/mm<sup>2</sup>) | -0.9 [-1.7,-0.1] | -1.8 [-3.5,-0.1] | 0.04 |

Values are model prediction ± 95%CI
P-values, which compare to no change, were calculated with mixed effect models with repeated measures with random participant intercept and slope and fixed effect of time
Abbreviations: DXA, dual-energy X-ray absorptiometry; aBMD, areal bone mineral density; QCT, quantitative computed tomography; vBMD, volumetric bone mineral density; HRpQCT, high-resolution peripheral quantitative computed tomography.

Postoperative changes in estimated bone strength were apparent at the tibia, although not at the radius (Table 3). Declines in failure load, stiffness, and apparent modulus were all worse in postmenopausal women (Figure 1h, 1i, and 1j). The predicted decline in failure load over 12 months was −5.7% in postmenopausal women vs. −1.7% in premenopausal women vs. +0.3 in men; the predicted decline in stiffness over 12 months was −5.5% in postmenopausal women vs. −0.9% in premenopausal women vs. +1.3% in men; and the predicated decline in apparent modulus over 12 months was −5.2% in postmenopausal women vs. −0.9% in premenopausal women vs. +1.3% in men.

### Comparison of participant characteristics and metabolic outcomes in the SG and RYGB cohort studies

We used data from our previous pre-post observational cohort study of adults undergoing RYGB (13), which employed an identical study design and measurements, to make careful comparisons of the skeletal effects of SG and RYGB. At preoperative baseline, characteristics were similar between cohorts. There were no differences in age, sex and menopausal distribution, race, baseline weight and BMI, calciotropic hormone levels, or FCA. There were more participants with diabetes in the RYGB cohort than in the SG cohort (40% vs. 19%, p = 0.03).

Postoperatively, percentage weight loss after SG approached, but was smaller than that after RYGB (−21.9% after SG vs. −25.1% after RYGB over 6 months; −28.8% after SG vs. −30.8% after RYGB over 12 months; p < 0.01 for procedure*time interaction in adjusted models). Decrease in intestinal fractional calcium absorption over 6 months, reported previously not only for RYGB (28) but also after SG (26), was worse after RYGB vs. SG (absolute mean decline of - 25.8% vs. −17.9%, p < 0.01 procedure*time interaction in adjusted models).

### Changes in skeletal parameters in comparison with RYGB

Increases in bone turnover marker levels were larger after RYGB than after SG. The bone resorption marker CTx increased by a median +276% (IQR +166% to +395%) over the 6 months after RYGB vs. by +156% (IQR +89% to +279%) after SG (p <0.01 for difference between procedures). The bone formation marker P1NP 6 months postoperatively increased by a median +112% (IQR +71% to +153%) after RYGB vs. by +58% (IQR +33% to +100%) after SG (p <0.01 for difference between procedures). Differences between procedures persisted even with adjustment for age, sex, menopausal status, and race.

By DXA, there was no evidence for differences between SG and RYGB in the extent of aBMD decline at the proximal femur or lumbar spine. However, decline in lumbar spine vBMD by QCT was larger after RYGB than SG (−7.8% after RYGB vs. −3.2% after SG over 12 months; Figure 2a). There was also a larger decline after RYGB than SG in total vBMD at the radius (Figure 2b). In terms of bone microarchitecture, at the radius, cortical thickness decreased more and trabecular area increased more after RYGB (Figures 2c and 2d). In contrast, there were larger changes after SG than RYGB in the trabecular compartment at the tibia, including larger decreases in trabecular number and larger increases in trabecular separation and trabecular heterogeneity (Figures 2e, 2f, 2g). There was no evidence for differences between surgical procedures in estimated strength parameter change. All models were adjusted for age, sex, menopausal status, race, preoperative weight, and diabetes status. To test whether the larger weight loss after RYGB explained the larger change after RYGB for spinal vBMD or the larger changes after RYGB at the radius for total vBMD, cortical thickness, or trabecular area, we entered weight change into the multivariable model. For spinal vBMD change, adjustment for weight change diminished the difference between surgical procedures, in that the point estimate decreased by 31% and the p-value for procedure*time was no longer statistically significant. In contrast, for radius total vBMD, cortical thickness, and trabecular area, point estimates changed minimally and the differences between RYGB and SG maintained statistical significance.

**Figure 2:**
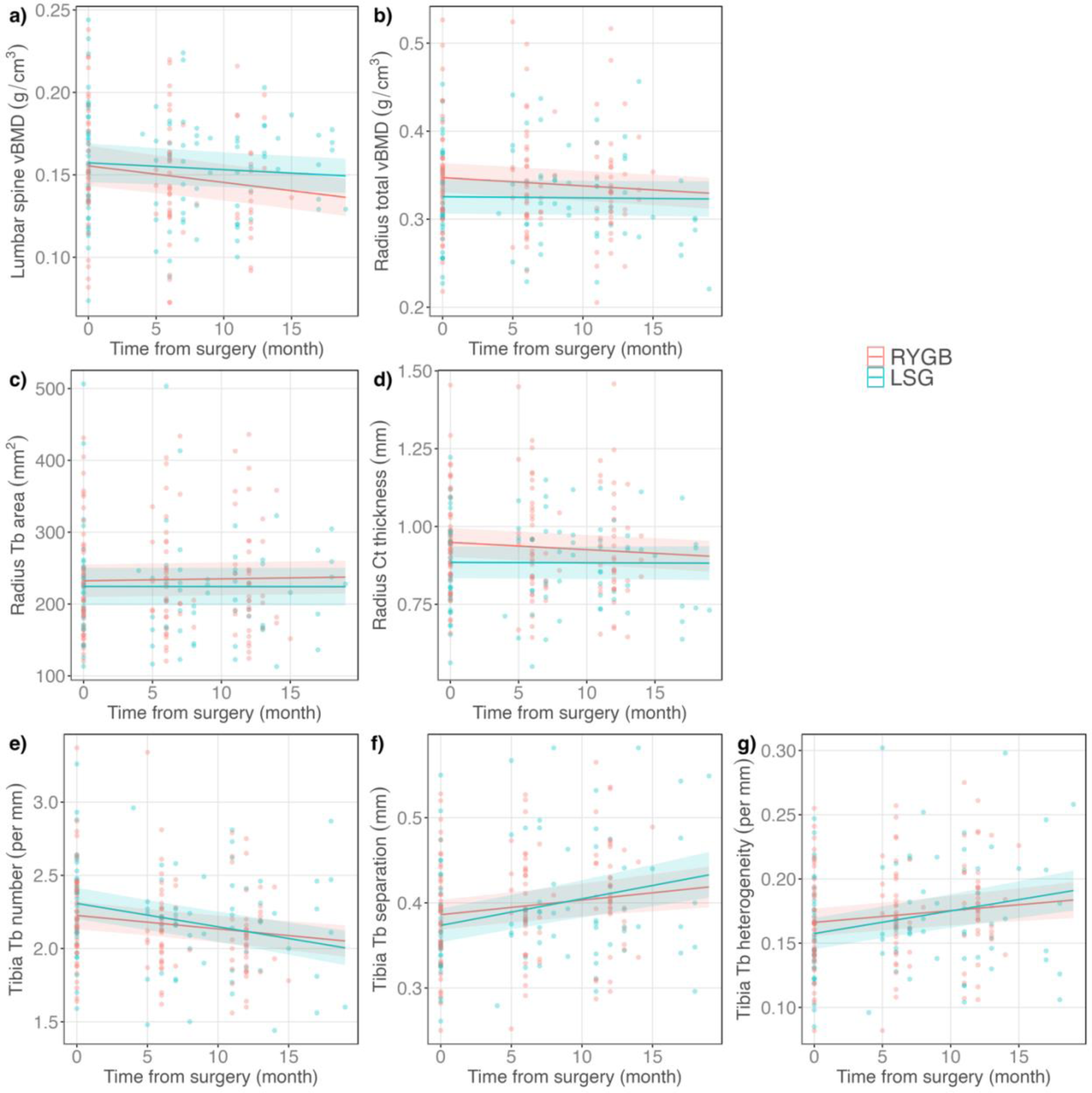
Changes in BMD, bone microarchitecture, and estimated strength that were significantly different by surgical procedure. Shaded areas represent the 95% confidence intervals.

## Discussion

This prospective cohort study of SG and skeletal health is the largest to date to examine axial and appendicular vBMD and appendicular bone microarchitecture and estimated strength. We found detrimental effects of SG on bone mass, structure, and strength that worsened progressively throughout the study duration (up to 20 months postoperatively). Our study is unique in its examination of the relative skeletal effects of SG by sex and menopausal status. Postmenopausal women are the population most burdened by lower bone mass and worse bone microstructure, and in our data this group experienced the largest declines in many skeletal health parameters after SG, including changes in BMD, bone microstructure, and estimated strength.

At the axial skeleton, postoperative declines in BMD occurred early and were substantial, particular at the proximal femur. The predicted 12-month percentage changes in aBMD were comparable to the magnitude of bone loss expected over the 3-4 years of fastest bone loss during the menopausal transition (45). Although the changes in lumbar spine vBMD by QCT overall did not reach statistical significance, a decline was observed in women that was offset by an increase in men. It is unclear why men had an increase in spine vBMD. One speculation is that men tend to have more degenerative disc disease, which may be a cause of spurious elevation not only for spine aBMD by DXA but also spine vBMD by QCT (46). In addition, in the setting of higher preoperative weight and waist circumference in men, greater preoperative abdominal soft tissue extension outside the CT scanner field of view could have result in more beam hardening artifact (47). Beam hardening could result in underestimation of vBMD at baseline and then an apparent increase in vBMD as weight is lost. Although QCT avoids the biases of DXA that stem from 2-dimensional, single-projection data acquisition, QCT assessments have nevertheless been shown to be influenced by obesity and weight loss (48).

There were also statistically significant declines in appendicular vBMD, albeit smaller in magnitude than at the axial sites. At the radius and tibia, both the trabecular and cortical compartments sustained detrimental effects, including decreases in trabecular number and increases in trabecular separation and heterogeneity and cortical pore size. The fact that these changes occurred at both the weight-bearing tibia and non-weight-bearing radius suggests at least in part a systematic nature of the skeletal effects of SG. However, the deterioration in bone density and microarchitecture was more pronounced at the weight-bearing skeleton. At the tibia, there were additional increases in trabecular area with decreases in cortical thickness at the tibia, consistent with endocortical resorption, and the impairment in bone density and microarchitecture translated into declines in estimated strength. The specificity for the tibia suggests that the mechanical unloading of weight loss may contribute in part to overall skeletal changes.

Postmenopausal women had worse declines than premenopausal women and men in some bone mass and microarchitectural parameters and in all estimated strength parameters at the tibia. This may reflect a heightened sensitivity of the already vulnerable postmenopausal skeleton—already vulnerable from age and sex hormone deficiency—to SG-induced bone metabolism abnormalities. The findings corroborate our previously reported finding that postmenopausal women were particularly impacted by skeletal effects of RYGB (13). If postmenopausal women are at highest risk for bariatric surgery-induced skeletal complications, there are implications for clinical care; it may be appropriate to target postmenopausal women with skeletal health screening, monitoring, and therapeutic interventions as they undergo RYGB or SG. Interventions have been studied: For example, a randomized controlled trial of a multipronged program of exercise, calcium, vitamin D, and protein supplementation was able to attenuate postoperative bone loss compared to no intervention in premenopausal women and men undergoing RYGB or SG (49). Ongoing clinical trials are now examining the safety and efficacy of osteoporosis pharmacologic interventions to reduce bariatric surgery-associated bone loss.

We were uniquely positioned to compare the skeletal effects of SG to those of RYGB, as we previously conducted a pre-post RYGB cohort study with identical study design and protocol (13). The two cohorts were remarkably similar in baseline characteristics, but we nevertheless adjusted our models for key baseline parameters. Increases in bone turnover marker levels were larger after RYGB than after SG. For DXA-assessed aBMD change, we did not detect differences between the surgical procedures. This finding is consistent with a recent meta-analysis of 14 studies on the effects of RYGB vs. SG on bone mineral density, which did not identify significant differences in aBMD change at the total hip, femoral neck, and lumbar spine (50). No prior studies have examined QCT or HRpQCT BMD, bone microstructure, or estimated strength between procedures. By QCT, we found that spinal vBMD decline was worse after RYGB than after SG. By HRpQCT, at the radius we demonstrated worse total vBMD decline and worse cortical thickness decline (with associated increase in trabecular area) after RYGB than after SG. Conversely, at the tibia, SG seemed to lead to worse microarchitectural changes in the trabecular compartment than RYGB. There were no differences in the decline in estimated bone strength between the two surgical procedures.

A number of potential factors may explain differential skeletal outcomes after the two surgical procedures. One such factor is extent of weight loss, which was not quite as large after SG as after RYGB. Our analyses suggest that the larger decline in spinal vBMD after RYGB compared to SG was explained by the greater weight loss experienced by the RYGB cohort. However, the larger declines at the radius for total vBMD and cortical thickness (with associated increase in trabecular area) after RYGB were not explained by greater weight loss after RYGB. Participants in both our cohorts underwent measurement of intestinal fractional calcium absorption, reported previously (26, 28), using a gold-standard dual stable isotope method. Decline in intestinal calcium absorption after SG, while marked, was less severe than after RYGB. This could contribute to the larger bone turnover marker increases and larger changes in select parameters at the radius after RYGB compared to SG. There are fewer potential explanations for our finding that SG seemed to lead to worse trabecular microarchitectural changes than RYGB at the tibia. In a published study comparing RYGB and SG over 12 months, there was an increase in bone marrow adiposity after SG but not RYGB (51), and one could speculate that an increase in marrow adiposity could lead to worsening trabecular microarchitecture.

Major strengths of our study include its prospective, longitudinal design, and the very comprehensive measurement of skeletal health using DXA, QCT, and HRpQCT as well as biochemical markers of bone turnover. Our cohort is unique as it included nearly 30% of postmenopausal women and nearly 25% of men, who are often excluded from or play a small role in SG research, because the majority of patients undergoing bariatric surgery are premenopausal women. Indeed, in a prior worldwide study from 2018, 73.7% of patients undergoing bariatric surgery were women with a median (IQR) age of 42 years (33–51) (17). Our study protocol included the careful, individualized supplementation of calcium and vitamin D in order to study the skeletal effects of SG in the setting of adherence to current standardized professional recommendations (52).

A limitation of our study is its modest duration; we did not determine the longer-term skeletal effects in our cohorts. The COVID19 pandemic impeded our ability to adhere to the protocol’s predefined timeline for study visits. However, we completed follow-up when we were able to do so, in fact exceeding our planned sample size, and we revised our statistical analysis plan to employ robust mixed-effect models taking into account the time to follow-up. Although we were able to make comparisons between SG and RYGB procedures using data from pre-post cohort studies that employed the same protocol and measurements and drew from the same bariatric surgery center, this was not a randomized controlled trial, nor was there a nonsurgical control group for full comparison. While our study is the largest to date to examine axial and appendicular vBMD and appendicular bone microarchitecture and strength, the sizes of the sex and menopause subgroups were modest. Future studies should enroll larger groups of postmenopausal women and men.

In conclusion, SG negatively impacts axial and appendicular BMD and appendicular bone microarchitecture and estimated strength. Increases in bone turnover marker levels and declines in some measures of bone mass and microarchitecture, while clinically and statistically significant after SG, are less severe after SG than after RYGB. For many other measures of bone mass, microarchitecture, and estimated strength, our data do not provide evidence for differences between the surgical procedures, or our data even suggest larger changes after SG. Postmenopausal women may be at highest risk of skeletal consequences after SG and RYGB, and thus targeted screening, monitoring, and interventions may be particularly appropriate for postmenopausal women undergoing SG. Further research should evaluate approaches to the prevention of long-term skeletal consequences of these otherwise beneficial metabolic procedures.

## Data Availability

All data produced in the present study are available upon reasonable request to the authors

## Acknowledgements

The authors thank Elliazar Enriquez, LVN for his facilitation of study recruitment; Aldric Chau for his work on QCT scan analysis; and Dolores Shoback, MD for her expert advice.

